# The effect of altruism on COVID-19 vaccination rates*

**DOI:** 10.1101/2022.06.15.22276430

**Authors:** Luis Á. Hierro, David Patiño, Pedro Atienza, Antonio J. Garzón, David Cantarero

**Affiliations:** University of Seville; University of Cantabria

**Keywords:** Altruism, vaccines, COVID-19, hesitancy, externality, herd immunity, public health

## Abstract

**Context:** After the emergence of the first vaccines against the COVID-19, public health authorities have promoted mass vaccination in order to achieve herd immunity and reduce the effects of the disease. Vaccination rates have differed between countries, depending on supply (availability of resources) and demand (altruism and resistance to vaccination) factors.

**Methods:** This work considers the hypothesis that individuals’ health altruism has been an important factor to explain the different levels of vaccination between countries, using the number of transplants as a proxy for altruism. Taking European Union’s countries to remove, as far as possible, supply factors that might affect vaccination, we carry out cross-sectional regressions for the most favorable date of the vaccination process (maximum vaccination speed) and for each month during the vaccination campaign.

**Results:** Our findings confirm that altruism has affected vaccination rates against the COVID-19. We find a direct relationship between transplants rates (proxy variable) and vaccination rates during periods in which the decision to be vaccinated depended on the individual’s choice, without supply restrictions. The results show that other demand factors have worked against vaccination: political polarization and belonging to the group of countries of the former Eastern bloc.

**Conclusions:** Altruism is a useful tool to define future vaccination strategies, since it favors the individuals’ awareness for vaccination.

## 1. Introduction

The fight against the COVID-19 pandemic has exposed inequalities in public health. At the beginning of the pandemic, these inequalities were manifested in clinical care and, later, in vaccination against SARS-Cov-2. The determining factor in a country’s vaccination capacity is income [1, 2]. High-income countries have not been financially constrained and have been able to access vaccines from the time they became available. In the first months of the vaccination campaign, they suffered temporary supply and logistics problems, but by the end of August 2021 vaccination had already reached rates of over 50% in most countries. However, the poorest countries –lacking the economic capacity to acquire vaccines– experienced very slow vaccination processes with very low vaccination rates [3]. This inequality in vaccination between rich and poor countries has triggered an important ethical debate [4]. However, despite the fact that rich countries have had vaccines –especially after the first months of vaccination– not all of them have followed the same vaccination patterns or have reached the same rates of vaccinated population. Previous studies show that gaps in vaccination levels are associated with disparities in income, educational level, gender and race, poverty status, etc. [5-7] and that citizens’ non-acceptance of the vaccine plays an important role [8, 9].

Social controversies surrounding vaccines have been common since the first vaccines were developed [10-16]. In recent decades, anti-vaccine movements have grown throughout much of the world [17, 18] such that, even before vaccines against SARS-Cov-2 became available, as early as 2020, surveys and studies into citizen willingness to get vaccinated were conducted [19-23]. Many such studies on pandemics have been carried out and there are already numerous works compiling their main results [24-27] or addressing specific groups such as health workers or students in the health area [28-31] or pregnant women [32]. These works reveal how people refuse to be vaccinated against SARS-Cov-2 for a variety of different reasons related to:

a. confidence in the vaccines themselves: their side effects or safety [33-39], their efficacy [39, 33, 40, 41], the need for it [40], and the development process behind these medicines [42]. Factors that favour trust or receptivity have also been analysed [43].
b. The development of pandemic and anti-pandemic measures: perception of the probability of contagion and severity of the disease [41, 44, 45, 38, 46]; the course of the pandemic [47, 48], the requirement of vaccination certificates for certain activities [49-51].
c. political issues, such as lack of trust in government [52, 20]; doubts about the vaccine authorization process [53, 54]; conspiracy theories [34, 44, 52], ideological positioning and political polarization [16, 34, 38, 41, 44, 55-61].

The cited studies have focused on factors that act against vaccination. However, there are factors that can work in favour of it. Indeed, the main characteristics of vaccines include their positive external effects; that is, the benefits they provide for the well-being of people other than those vaccinated (social benefits). When an individual is vaccinated, two positive external effects occur: a direct external effect, associated with the reduced likelihood of infecting people with whom we interact –especially direct family members– and a collective external effect, which occurs as a result of our vaccination reducing the general probability of contagion. This general probability of contagion depends inversely on the percentage of people vaccinated, although in a nonlinear way. When the percentage of those vaccinated is small, the external effects are also small. Positive external effects grow more than proportionally as the percentage of vaccinated people grows. When a certain threshold is reached in the proportion of immune individuals, the incidence of infection begins to decrease. At that moment, what is known as herd immunity is reached. [62]. Acquiring herd immunity is the fundamental instrument of public health in a pandemic situation while there is no drug available to mitigate or eliminate the effects of the disease [63-66].

If we use the term *health altruism* to refer to the assessment the individual makes of the social benefits of being vaccinated, then the greater the population’s health altruism, the higher a country’s vaccination rate must be and, therefore, the greater the possibility of achieving herd immunity. This work aims to pinpoint this effect by testing the hypothesis that social altruism favours vaccination against COVID-19 and proves decisive vis-à-vis the existence of different vaccination rates. For this purpose, the number of organ transplants is used as a proxy for health altruism, since the concept of altruism forms the basis of transplant ethics [67]. This means that we can consider transplants as a real manifestation of individuals’ will to contribute to the health of others and, therefore, of altruism in health matters.

The works published to date have not attempted to pinpoint the influence of health altruism on vaccination rates against COVID-19, such that there are no previous references or data from surveys. To test the hypothesis, we performed a cross-sectional analysis, taking vaccination data from European Union countries. This selection is due to the fact that the EU has centrally managed the purchase of vaccines for all EU countries and has distributed them equally, substantially reducing the influence of supply factors related to the accessibility and availability of the vaccine.

Our work shows that during the first phase of vaccination when supply constraints were in operation –that is, when there were insufficient vaccines available and the shortage conditioned vaccination– health altruism had no impact. However, when supply restrictions disappeared and individuals’ desire to be vaccinated became a factor, health altruism did come into play as an important variable to explain countries’ different vaccination. Likewise, this paper shows that other demand factors negatively affect vaccination: being a former Eastern European bloc country, and the level of political polarization.

The structure of the work is as follows: in section 2, we explain the methodology used and the data sources. In section 3, we show the results obtained, while in section 4 we comment on the results, analysing their limitations and implications. In section 5, we present the conclusions.

## 2. Methodology and data

The vaccination process is determined not only by subjects’ willingness to be vaccinated, which manifests itself in the demand for vaccination, but also by the availability of the vaccine; that is, by supply. When the government assumes vaccination, there are basically three supply factors: a) vaccines are available on the market, b) the government can pay for vaccines, and c) the government has the logistics and health system required to carry out mass vaccination. Factors b) and c) are related to the country’s economic capacity, while factor a) –the existence of available vaccines– is related to the organization of a global manufacturing system.

### 2.1. Geographical scope

To minimize the impact of factors b) and c), we chose to limit the study to a restricted geographical area: the European Union. The European Union assumed the acquisition and distribution of vaccines in order to achieve equitable distribution and reduce the effects of competition between countries on the price and availability of vaccines. Furthermore, the EU has also financed governments to cover the costs of the pandemic, which has avoided substantial differences in distribution logistics. The result of this European intervention is that the influence of supply factors b) and c) is almost eliminated if we select this particular geographical area.

### 2.2. Timeframe

For its part, factor a) –scarcity of vaccines– is related to the moment. The more advanced the vaccination campaign, the smaller the effect of the vaccine shortage will be. Therefore, if we wait until the end of the vaccination process, we eliminate the effect of the shortage. However, this implies missing out on relevant information for public health authorities in terms of influencing the vaccination process, such that we must look for another alternative.

The problem is that choosing any other day implies a bias, which in this work we try to eliminate by using two methods: I) The first involves taking the day on which the vaccination speed is at its highest for each country; II) while the second involves making a cross-section at the end of each completed month since the start of vaccination in the European Union.

For Method I), the concept of vaccination speed or average daily number of vaccinated people is used; that is, the ratio between the total number of those vaccinated and the number of days to have elapsed since the start of vaccination, choosing when this ratio is at its maximum for each country. By making this choice, we minimize any supply restriction, although it does raise the problem that we fail to consider the same date for all countries.

Method II) involves estimating the model month by month and checking how the significance of the coefficients of the different variables evolves. In principle, this method allows us to know the effect of altruism with and without supply restrictions as well as the time consistency of the results, although the problem is that it is influenced by changes in health policy and by the course of the disease and the virus.

### 2.3. Model specification

The specification to be estimated is given by:

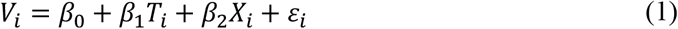

where *V*_*i*_ indicates the percentage of the population fully vaccinated in country i on the corresponding date, and *T*_*i*_ is the representative variable of health altruism in country i. Since we do not have any type of survey that can give us an indicator of this altruism, we use the number of organ transplants in each country as a proxy, since they are the revealed expression of altruism at the highest level in health. Additionally, the vector *X*_*i*_ includes two demand variables that we introduce as control variables: political polarization, due to the relationship between antivaccine movements and political extremisms; and a dummy that identifies the country’s former membership of the Eastern bloc, in order to capture possible public sector distrust in those countries. As explained above, this specification is estimated using data on the day of maximum vaccination speed for each country in Method I and data of vaccination in each month in Method II.

### 2.4. Data Sources

The data sources for the variables included in the econometric specification are shown in the following Table 1.

**Table 1.** Variables used in the specification and data sources.

| Variable | Description of variables | Source |
| --- | --- | --- |
| Vaccination rate | Fully vaccinated people in % of the total population. | Ritchie, (2020) [68] |
| Organ transplants and donations | Total number of transplants or donations of any kind in % of the total population. | Global Observatory on Donation and Transplantation (GODT) (http://www.transplant-observatory.org/). This constitutes the most complete source of data worldwide on donations and transplants, information for which comes from the national organizations coordinated by the WHO (Data from the year 2020) |
| Political polarization index | Aggregation of four polarization indices: ideological and affective of the masses and ideological and affective of the elite. | Van der Veen (2021) [69]<br>The database is available online at: https://drive.google.com/drive/folders/1sErZ3lb-Z3eEr_zXqJ68PWnI-ksVxVxU (The latest available data has been taken: 2020) |

Source: authors' own compilation.

For those countries which, during the period studied, published vaccination data less frequently than on a daily basis or did not publish data at weekends, as well as for the days on which a country published no data, we linearly interpolate between data from the previous day and the following day in order to obtain daily frequency time series for each country without any missing values. This solution is impossible to apply to Bulgaria, since during a large part of the period it reported its data with a very low frequency, reaching periods of over three weeks without reporting new data. Given the impossibility of linear interpolation, we chose to exclude Bulgaria from the study. Finally, for Portugal and Cyprus no data was given for April, such that the level of vaccination on the last day published is considered.

## 3. Results

### 3.1. Influence of health altruism on vaccination against COVID-19. Method I

To apply Method I, we take 8 December 2020 –the day on which vaccination began in the EU– as the starting point for the sample, specifically in Denmark. We take the same day for all EU countries since, when buying vaccines at a European level, the availability of vaccines is simultaneous in all countries, such that differences in the starting day of vaccination are unrelated to the provision of vaccines. Table 2 collects the data corresponding to the date of maximum vaccination rate.

**Table 2.** Vaccination data against COVID-19 corresponding to the day of maximum vaccination speed in each European Union country.

| Country | Day of maximum vaccination speed | Number of days since 12/08/2020 | Total number of those vaccinated on the day of maximum speed (number of people) | Vaccination level on the day of maximum speed (% people over total) | Maximum vaccination speed (number of people / number of days) | Maximum vaccination rate (% of the total / number of days) |
| --- | --- | --- | --- | --- | --- | --- |
| Austria | 02/07/2021 | 207 | 4,971,228 | 54.97 | 24,015.59 | 0.2656 |
| Belgium | 03/07/2021 | 208 | 7,470,771 | 64.22 | 35,917.17 | 0.3088 |
| Check Republic | 25/06/2021 | 200 | 4,986,583 | 46.50 | 24,932.92 | 0.2325 |
| Croatia | 11/06/2021 | 186 | 1,401,647 | 34.34 | 7,535.74 | 0.1846 |
| Cyprus | 17/06/2021 | 192 | 443,121 | 49.46 | 2,307.92 | 0.2576 |
| Denmark | 29/07/2021 | 234 | 4,149,738 | 71.38 | 17,733.92 | 0.3051 |
| Estonia | 18/06/2021 | 193 | 540,086 | 40.76 | 2,798.37 | 0.2111 |
| Finland | 04/07/2021 | 209 | 3,380,819 | 60.93 | 16,176.17 | 0.2915 |
| France | 13/08/2021 | 249 | 46,673,270 | 69.23 | 187,442.85 | 0.2780 |
| Germany | 09/07/2021 | 214 | 49,023,479 | 58.43 | 229,081.68 | 0.2730 |
| Greece | 23/07/2021 | 228 | 5,578,596 | 53.79 | 24,467.53 | 0.2359 |
| Hungary | 22/05/2021 | 166 | 5,005,682 | 51.96 | 30,154.71 | 0.3130 |
| Ireland | 04/08/2021 | 240 | 3,460,704 | 69.45 | 14,419.60 | 0.2894 |
| Italy | 02/07/2021 | 207 | 34,862,781 | 57.75 | 168,419.23 | 0.2790 |
| Latvia | 17/11/2021 | 345 | 1,250,025 | 66.96 | 3,623.26 | 0.1941 |
| Lithuania | 20/08/2021 | 256 | 1,607,299 | 59.75 | 6,278.51 | 0.2334 |
| Luxemburg | 17/07/2021 | 222 | 384,752 | 60.61 | 1,733.12 | 0.2730 |
| Malta | 25/05/2021 | 169 | 310,403 | 60.14 | 1,836.70 | 0.3559 |
| Netherlands | 10/07/2021 | 215 | 11,691,199 | 68.08 | 54,377.67 | 0.3166 |
| Poland | 18/06/2021 | 193 | 16,045,779 | 42.45 | 83,138.75 | 0.2200 |
| Portugal | 29/08/2021 | 265 | 8,610,175 | 84.68 | 32,491.23 | 0.3195 |
| Romania | 22/05/2021 | 166 | 4,104,686 | 21.46 | 24,727.02 | 0.1293 |
| Slovakia | 31/05/2021 | 175 | 1,811,015 | 33.16 | 10,348.66 | 0.1895 |
| Slovenia | 18/06/2021 | 193 | 814,831 | 39.20 | 4,221.92 | 0.2031 |
| Spain | 19/08/2021 | 255 | 35,442,173 | 75.82 | 138,988.91 | 0.2973 |
| Sweden | 25/07/2021 | 230 | 6,253,678 | 61.55 | 27,189.90 | 0.2676 |

Source: authors' own elaboration based on data from Mathieu, (2021) [70].

Finally, Table 3 collects the estimated specification. As can be seen, the results show that the representative variable of health altruism positively influences the vaccination rate, thereby confirming our hypothesis. Likewise, the other two demand variables –political polarization, and the country’s membership of the Eastern bloc– work against vaccination and are statistically significant.

**Table 3.** Estimated influence of health altruism on the vaccination rate of European Union countries corresponding to the day of maximum vaccination rate in each European Union country.

| **Demand variables** | **Estimated coefficient** |
| --- | --- |
| Transplants | 637,872**<br>(243,583) |
| Polarization Index | -6.765*<br>(3.428) |
| Former Eastern Bloc country | -19.09***<br>(4.574) |
| Constant | 84.76***<br>(14.00) |
| Observations | 26 |
| R-square | 0.595 |

Source: original elaboration based on data from Mathieu, (2021) [70]. Note: robust standard errors in parentheses.
\*\*\* p<0.01, \*\* p<0.05, \* p<0.1

### 3.2. Influence of health altruism on vaccination against COVID-19. Method II

To apply Method II, we collected data on vaccination levels on the seventh of every month, starting on 8 December 2020. This was the day vaccination began in Denmark, and we conducted a periodic cross-sectional study for those days, until the last available day, considering the same demand variables.

Table 4 shows the results obtained. As can be seen, during the first phase of vaccination –when supply constraints are in operation– demand variables have no effect. However, when supply restrictions disappear and individual will to be vaccinated begins to weigh more, these variables become relevant.

**Table 4.** Estimation of the impact of health altruism on the vaccination rate of European Union countries corresponding to each month since the start on 8 December 2020.

| Date | (1)<br>7/1/21 | (2)<br>7/2/21 | (3)<br>7/3/21 | (4)<br>7/4/21 | (5)<br>7/5/21 | (6)<br>7/6/21 | (7)<br>7/7/21 | (8)<br>7/8/21 |
| --- | --- | --- | --- | --- | --- | --- | --- | --- |
| Transplants | 13,808<br>(9,009) | -19,574<br>(17,617) | -13,254<br>(39,004) | -50,554<br>(117,786) | -48,055<br>(175,916) | 12,386<br>(201,968) | 243,583<br>(195,321) | 440,820**<br>(189,993) |
| Polarization Index | -0.160<br>(0.197) | -0.396<br>(0.424) | -1.073<br>(0.944) | -2.339<br>(2.840) | -2.573<br>(3.811) | -2.331<br>(3.998) | -4.206<br>(3.267) | -5.857*<br>(3.195) |
| Former Eastern Bloc | 0.122<br>(0.152) | -0.392<br>(0.328) | -0.651<br>(0.696) | -0.832<br>(1.896) | -4.146<br>(2.627) | -7.637**<br>(2.765) | -15.91***<br>(2.796) | -19.86***<br>(2.796) |
| Constant | 0.879<br>(0.869) | 5.263**<br>(2.265) | 12.65**<br>(4.905) | 27.19*<br>(14.25) | 42.45**<br>(18.52) | 55.25***<br>(19.15) | 73.93***<br>(14.39) | 87.01***<br>(14.14) |
| Observations | 26 | 26 | 26 | 26 | 26 | 26 | 26 | 26 |
| R-square | 0.091 | 0.181 | 0.141 | 0.087 | 0.144 | 0.274 | 0.641 | 0.756 |

| Date | (9)<br>7/9/21 | (10)<br>7/10/21 | (11)<br>7/11/21 | (12)<br>7/12/21 | (13)<br>7/1/22 | (14)<br>7/2/22 | (15)<br>7/3/22 | (16)<br>7/4/22 |
| --- | --- | --- | --- | --- | --- | --- | --- | --- |
| Transplants | 599,948***<br>(201,950) | 600,481***<br>(203,854) | 569,776**<br>(207,967) | 545,537**<br>(209,018) | 521,730**<br>(206,180) | 468,392**<br>(203,250) | 436,847**<br>(190,999) | 414,240**<br>(177,106) |
| Polarization Index | -6.154*<br>(3.423) | -5.878<br>(3.454) | -6.368*<br>(3.400) | -6.114*<br>(3.197) | -6.164*<br>(3.161) | -5.768*<br>(3.252) | -5.596*<br>(3.207) | -5.434*<br>(3.121) |
| Former Eastern Bloc | -21.20***<br>(3.044) | -20.55***<br>(2.996) | -18.94***<br>(3.127) | -17.87***<br>(2.986) | -18.31***<br>(3.006) | -19.29***<br>(2.998) | -19.30***<br>(2.921) | -19.20***<br>(2.836) |
| Constant | 90.50***<br>(15.04) | 91.08***<br>(15.14) | 95.31***<br>(14.74) | 96.33***<br>(13.57) | 98.83***<br>(13.49) | 99.74***<br>(14.27) | 99.81***<br>(14.39) | 99.61***<br>(14.30) |
| Observations | 26 | 26 | 26 | 26 | 26 | 26 | 26 | 26 |
| R-square | 0.755 | 0.751 | 0.720 | 0.719 | 0.720 | 0.726 | 0.730 | 0.737 |

Source: original elaboration based on data from Mathieu, (2021) [70]. Note: robust standard errors in parentheses. \*\*\* p<0.01, \*\* p<0.05, \* p<0.1.

Estimates by month throughout the period in which vaccination took place show a positive relationship between health altruism and vaccination rate, starting from the period in which restrictions on the supply of vaccines began to disappear and the vaccine began to be fully accessible (eighth month of vaccination). In the same period, the estimates capture the influence of political polarization and being a former Soviet bloc country. In the latter case, the negative relationship appeared a couple of months earlier. As can be seen in the table, the results are robust throughout the period, both in the value of the estimated coefficients and in their statistical significance.

Alternatively, as a robustness check, we estimate both methods using organ donations instead of transplants. The results are similar and are included in Appendix B of Supplementary Information.

## 4. Discussion

The results obtained show that individuals’ altruism, measured through a proxy variable such as transplants per capita, favours vaccination against COVID-19. The results are robust and are supported by the two methodologies used.

### 4.1. The role of health altruism in vaccination

Countries that traditionally show a more committed attitude towards the health of others –that is, countries which value the external effects of their behaviour and demonstrate this through a greater number of transplants– have achieved higher rates of vaccination against COVID-19. When we take the vaccination rate at the time of the highest vaccination speed, the value of the coefficient is greater than 600 and when we estimate month by month, the significance of the coefficient is maintained over time from the eighth month of vaccination, although the value of its coefficient begins to decrease from the eleventh month. This evolution of the estimated coefficient could be related to: a) the loss of altruism weight as vaccination progresses and people are already vaccinated; b) the course of the pandemic, since the appearance of the omicron variant caused the vaccination processes to accelerate again due to the selfishness of those who were unvaccinated; and c) because European governments began to require vaccination passports for work and leisure [50]. In this sense, further research would be needed.

### 4.2. The impact of other demand and supply factors

On the opposite side of altruism, there are the other two demand factors included in the specification that work against vaccination. On the one hand, the dummy variable representing the country’s membership of the Eastern bloc shows that they have vaccination rates that are around 19% lower, which bears out previous results [71] that already anticipated the negative effect of this condition, as a consequence of elderly people’s distrust of public policies.

On the other hand, political polarization –also related to political mistrust– eventually manifesting itself in antivaccine positions and lower vaccination rates, which is also captured by our estimation using the two methods [72]. The relationship between electoral support for populist political parties and rejection of vaccination was already evident before the COVID-19 pandemic, and shows how political populism and vaccine rejection are driven by similar dynamics: deep mistrust towards elites and experts. Later, during the pandemic, political extremism reduced the population’s willingness to comply with social distancing measures [60] and caused rejection of vaccination [16], a result that our work also confirms.

As regards supply factors, as expected, with method I), by assessing vaccination at the time of maximum speed, or average daily vaccination, we were able to isolate the effects of these supply factors, as was our intention. However, as we observed with method II) supply factors would have had an impact until the sixth month of vaccination, and the estimate does not stabilize until the eighth month of vaccination. We must understand that this is because, in that period, the European Union still lacked sufficient vaccines, and that it was therefore the supply shortage that conditioned vaccination.

### 4.3. Limitations

The work has limitations that must be taken into account. First of all, we must consider the restricted geographical area. We only have 26 observations in each estimate, which has limited the possibility of expanding the number of variables. The negative effect of this reduced number of observations has been offset by removing the effect of supply factors, yet any replication of the work for larger geographical areas will require these variables to be introduced so as to control for economic and logistical constraints in countries with lower income per capita.

In addition, the work does not control the impact of the course of the disease itself (new variants of SARS-Cov 2, waves of contagion…) or the measures adopted to control the spread of the disease. Both should impact how vaccination progressed, and it is therefore reasonable to assume that they would alter the relevance of altruism in vaccination. In fact, the variation in the coefficients estimated from month 12 for the proxy variable is very likely related to this issue.

### 4.4. Implications

Important implications emerge from the results. First, we observe that during the period of supply shortage, the fundamental objective must be to eliminate this shortage, since the other factors are not relevant for vaccination if production is not capable of meeting demand. Once supply constraints begin to ease, demand factors come into play and altruism favours vaccination. For this reason, health authorities should promote vaccination, for example, through advertising campaigns to raise public awareness that influence health altruism and spread the benefits of vaccination for the health of people around us.

In addition, given that countries where health altruism is more developed have an initial advantage in terms of vaccination processes, health authorities must invest preventively and systematically to promote such health altruism. In particular, governments should value fostering health altruism in education, since this can yield high social returns in the long term when we are faced with new pandemics.

Another implication of the results is the need to prevent rejection of vaccination from radical political positions. Governments must be aware that political radicalism operates against vaccination and they must work preventively by drawing up contingency plans to reduce anti-vaccine reaction in radicalized countries. In this sense, it may be important to promote agreements between parties vis-à-vis leaving vaccines out of the political dispute and establishing regulations that make vaccination mandatory, or that sanction non-vaccination (prohibiting access to workplaces and the hospitality/catering sector for unvaccinated people, or mandatory vaccination passports). These actions must be planned in advance and must take effect at a time when there is no shortage of vaccines and when the speed of vaccination continues to increase.

This last recommendation –that governments should focus their efforts on the first phase of vaccination– is deduced from the relationship of the final vaccination rate with the maximum vaccination speed and the time elapsed until it is reached. As can be seen in the estimate shown in Table 5, lengthening the period until reaching maximum speed and increasing the latter are decisive in terms of achieving a high level of final vaccination. Both factors explain almost 90% of the value of the final vaccination rate.

**Table 5.** Relationship between the maximum vaccination speed and the days required to reach it with the final level of vaccination by country.

| Variable | Coefficient |
| --- | --- |
| Constant | -2.1046<br>(4.4011) |
| Days to maximum speed | 0.1163***<br>(0.0267) |
| Maximum vaccination speed | 195.4141***<br>(17.2801) |
| Observations | 26 |
| R-square | 0.8845 |

Source: own elaboration. Notes: OLS regression. Robust standard errors in parentheses. \*\*\* p<0.01, \*\* p<0.05, \* p<0.1.

Finally, it is necessary to reflect on Eastern European countries and their particularity in terms of vaccination. The effects of low vaccination rates against COVID-19 manifest themselves in an excess mortality of close to 30% [73], coinciding with vaccination rates which, as we have seen, were around 19% lower. This situation highlights the need to influence the study of these countries and to adopt measures to reverse this mistrust that proves so detrimental to public health.

## 5. Conclusions

There are factors that hinder mass vaccination of the population against COVID-19 and that lead to differences in vaccination rates between countries. These factors are, firstly, supply factors since vaccination is affected by countries’ economic capacity to acquire vaccines and the logistical problems involved in a mass vaccination campaign. On the demand side, the main factor that hinders achieving herd immunity is the population’s refusal to be vaccinated. This latter aspect has been studied in the literature, as described in this work.

However, there are demand factors that can favour vaccination, which leads to differences in vaccination rates between countries and which therefore has implications for public health policy. This article tests the hypothesis that health altruism has a positive impact on population vaccination. We represent altruism in health through a proxy variable –organ transplants– to reflect the greater acceptance of vaccination when individuals assume the positive external effects for their direct relatives and for society in general, by helping to achieve herd immunity.

The results obtained support the hypothesis that health altruism is an explanatory factor of demand, and positively influences countries’ vaccination rates. Likewise, other demand factors that work against vaccination have been pinpointed, such as the country’s political polarization or being a former Eastern Bloc country. As a consequence of these results –and bearing in mind that pandemics such as COVID-19 might happen again– the study concludes that governments must plan actions to favour vaccination by systematically promoting health altruism in the long term. These actions guarantee a good starting point for possible future pandemic contexts. In addition, it is also necessary to plan positive and negative incentive processes for vaccination in a pandemic –focusing on the first phase of vaccination– so as to reduce the negative impact of the factors that act against vaccination.

## Data Availability

All data produced in the present study are available upon reasonable request to the authors

## Appendix A.

### Evolution of vaccination across European Union countries

**Figure A1.**
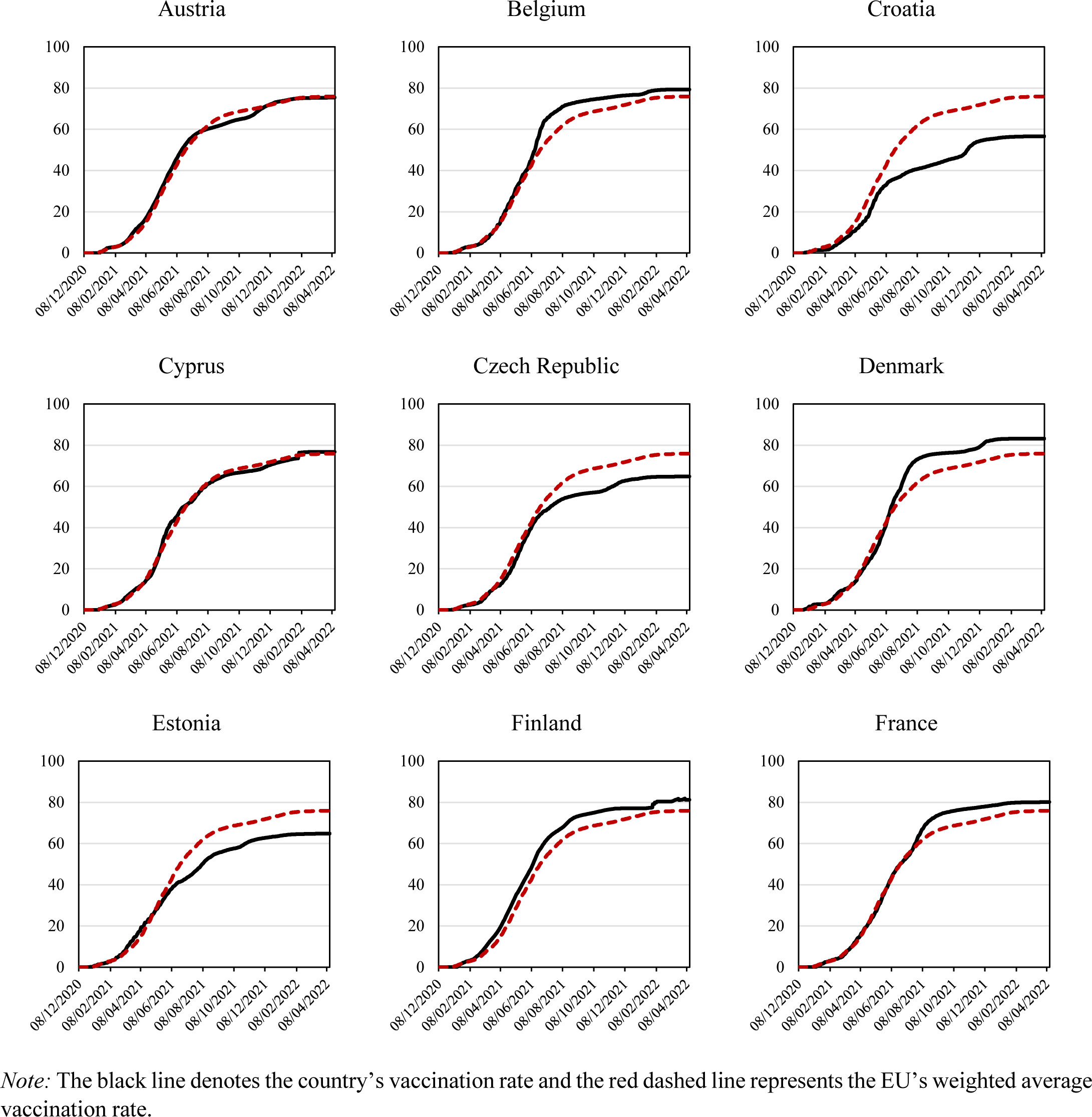
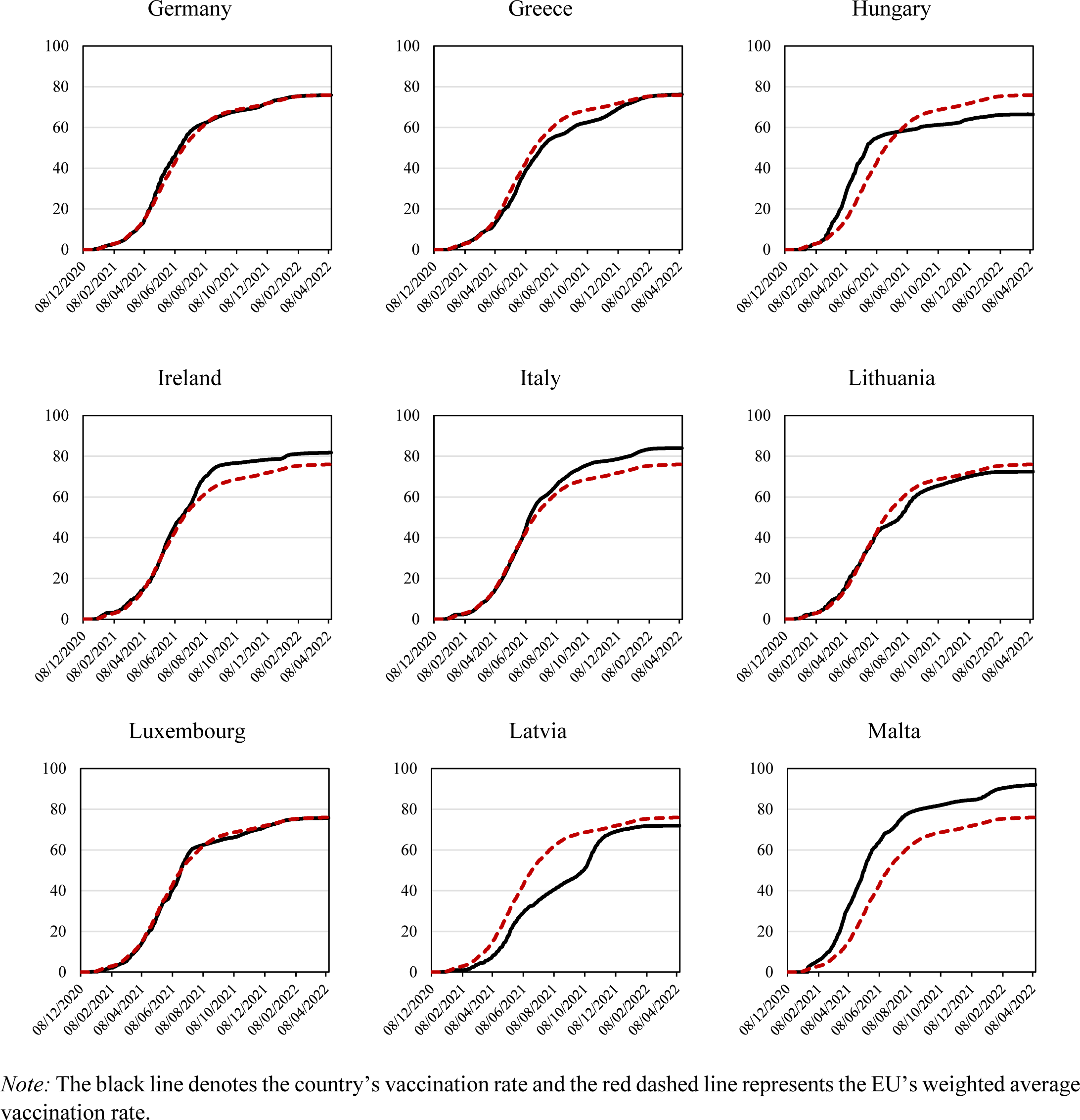
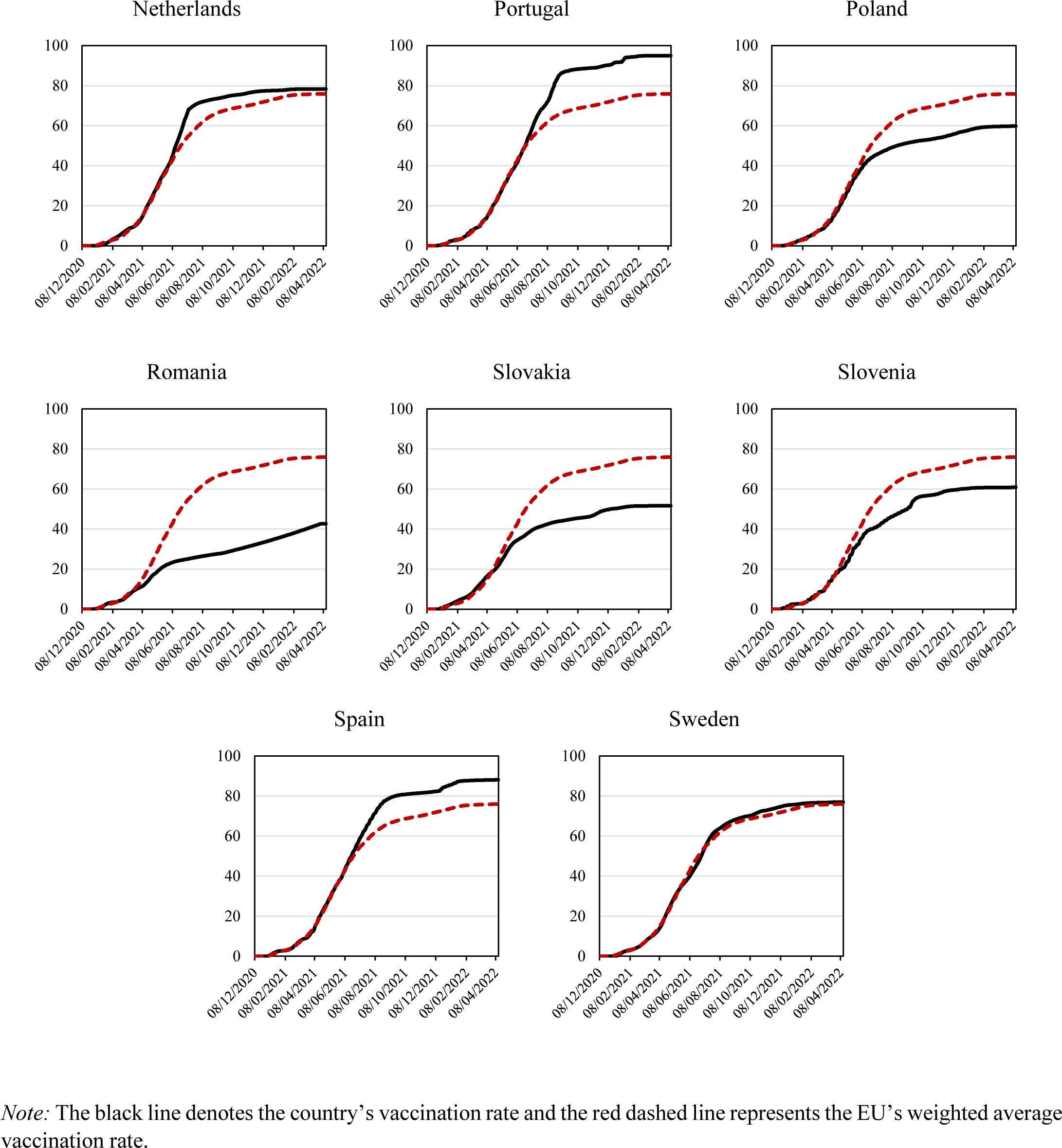
Evolution of the vaccination rate by country.

## Appendix B.

### Results using organ donations as proxy for altruism

**Table B1.** Estimated influence of health altruism on the vaccination rate of European Union countries corresponding to the day of maximum vaccination rate in each country.

| Demand variables | Estimated coefficients |
| --- | --- |
| Donations | 564,745**<br>(221,907) |
| Polarization Index | -6.700*<br>(3.545) |
| Former Eastern Bloc | -19.19***<br>(4.611) |
| Constant | 85.15***<br>(14.57) |
| Observations | 26 |
| R-square | 0.588 |

Source: original elaboration based on data from Mathieu, (2021) [70]. Note: robust standard errors in parentheses.
\*\*\* p<0.01, \*\* p<0.05, \* p<0.1

**Table B2.** Estimation of the incidence of health altruism in the vaccination rate of the countries of the European Union corresponding to each month since the start on December 8, 2020.

| Date | (1)<br>7/1/21 | (2)<br>7/2/21 | (3)<br>7/3/21 | (4)<br>7/4/21 | (5)<br>7/5/21 | (6)<br>7/6/21 | (7)<br>7/7/21 | (8)<br>7/8/21 |
| --- | --- | --- | --- | --- | --- | --- | --- | --- |
| Donations | 13,402<br>(8,345) | -18,220<br>(15,754) | -12,703<br>(34,770) | -50,668<br>(105,774) | -52,442<br>(159,546) | 3,871<br>(183,466) | 205,373<br>(180,988) | 381,946**<br>(178,021) |
| Polarization Index | -0.166<br>(0.197) | -0.393<br>(0.426) | -1.069<br>(0.950) | -2.308<br>(2.855) | -2.517<br>(3.831) | -2.286<br>(4.019) | -4.118<br>(3.286) | -5.761*<br>(3.248) |
| Former Eastern Bloc | 0.121<br>(0.151) | -0.390<br>(0.327) | -0.650<br>(0.695) | -0.832<br>(1.892) | -4.151<br>(2.625) | -7.648**<br>(2.774) | -15.96***<br>(2.824) | -19.94***<br>(2.844) |
| Constant | 0.900<br>(0.872) | 5.241**<br>(2.260) | 12.63**<br>(4.901) | 27.09*<br>(14.24) | 42.31**<br>(18.53) | 55.18***<br>(19.16) | 73.96***<br>(14.37) | 87.19***<br>(14.25) |
| Observations | 26 | 26 | 26 | 26 | 26 | 26 | 26 | 26 |
| R-square | 0.099 | 0.182 | 0.142 | 0.088 | 0.146 | 0.274 | 0.637 | 0.749 |

| Date | (9)<br>7/9/21 | (10)<br>7/10/21 | (11)<br>7/11/21 | (12)<br>7/12/21 | (13)<br>7/1/22 | (14)<br>7/2/22 | (15)<br>7/3/22 | (16)<br>7/4/21 |
| --- | --- | --- | --- | --- | --- | --- | --- | --- |
| Donations | 526,802**<br>(189,287) | 534,366***<br>(188,959) | 505,526**<br>(192,131) | 487,705**<br>(191,695) | 469,353**<br>(187,386) | 421,518**<br>(184,499) | 393,017**<br>(173,367) | 372,462**<br>(160,792) |
| Polarization Index | -6.066*<br>(3.493) | -5.834<br>(3.515) | -6.316*<br>(3.462) | -6.087*<br>(3.251) | -6.156*<br>(3.211) | -5.762*<br>(3.287) | -5.590*<br>(3.238) | -5.427*<br>(3.150) |
| Former Eastern Bloc | -21.29***<br>(3.111) | -20.64***<br>(3.040) | -19.02***<br>(3.178) | -17.94***<br>(3.035) | -18.38***<br>(3.050) | -19.35***<br>(3.037) | -19.36***<br>(2.956) | -19.25***<br>(2.868) |
| Constant | 90.82***<br>(15.25) | 91.47***<br>(15.34) | 95.67***<br>(14.94) | 96.72***<br>(13.77) | 99.23***<br>(13.69) | 100.1***<br>(14.39) | 100.1***<br>(14.48) | 99.93***<br>(14.37) |
| Observations | 26 | 26 | 26 | 26 | 26 | 26 | 26 | 26 |
| R-square | 0.746 | 0.745 | 0.714 | 0.714 | 0.717 | 0.723 | 0.728 | 0.735 |

Source: Original elaboration based on data from Mathieu, (2021) [70]. Note: robust standard errors in parentheses. \*\*\* p<0.01, \*\* p<0.05, \* p<0.1.

## References

1. Duan Y, Shi J, Wang Z, et al. Disparities in COVID-19 vaccination among low, middle and high-income countries: the mediating role of vaccination policy. Vaccines. 2021;9(8):905. https://doi.org/10.3390/vaccines9080905.

2. Roghani A, Panahi S. The global distribution of COVID-19 vaccine: The role of macro-socioeconomics measures. 2021. https://doi.org/10.1101/2021.02.09.21251436. Accessed 18 April 2022.

3. Padma TV. COVID-19 vaccines to reach poorest countries in 2023 — despite recent pledges. Nature. 2021;595:342–343. https://doi.org/10.1038/d41586-021-01762-w.

4. Jecker NS, Wightman AG, Diekema DS. Vaccine ethics: an ethical framework for global distribution of COVID-19 vaccines. Journal of Medical Ethics. 2021;47(5):308–317. http://dx.doi.org/10.1136/medethics-2020-107036.

5. Bauer C, Zhang K, Lee M, et al. Real-time geospatial analysis identifies gaps in COVID-19 vaccination in a minority population. Scientific Reports. 2021;11(1):1–6. https://doi.org/10.1038/s41598-021-97416-y.

6. Ishimaru T, Okawara M, Ando H, et al. Gender differences in the determinants of willingness to get the COVID-19 vaccine among the working-age population in Japan. Human Vaccines & Immunotherapeutics. 2021;17(11):3975–3981. https://doi.org/10.1080/21645515.2021.1947098.

7. Hughes MM, Wang A, Grossman M K, et al. County-level COVID-19 vaccination coverage and social vulnerability—United States, December 14, 2020–March 1, 2021. Morbidity and Mortality Weekly Report. 2021;70(12):431–436. https://dx.doi.org/10.15585%2Fmmwr.mm7012e1.

8. Liu R, Li GM. Hesitancy in the time of coronavirus: Temporal, spatial, and sociodemographic variations in COVID-19 vaccine hesitancy. SSM-population health. 2021;15:100896. https://doi.org/10.1016/j.ssmph.2021.100896.

9. Niño MD, Hearne BN, Cai T. Trajectories of COVID-19 vaccine intentions among US adults: The role of race and ethnicity. SSM-population health. 2021;15:100824. https://doi.org/10.1016/j.ssmph.2021.100824.

10. Nelson M, Rogers J. The Right To Die? Anti-Vaccination Activity And The 1874 Smallpox Epidemic In Stockholm. Social History of Medicine. 1992;5(3):369–388.

11. Porter D, Porter R. The politics of prevention: anti-vaccinationism and public health in nineteenth-century England. Medical history. 1988;32(3):231–252. https://doi.org/10.1017/S0025727300048225.

12. National Anti-Compulsory-Vaccination League (CHELTENHAM). Think for yourselves. 3rd rev. ed. Cheltenham; 1876.

13. Scarpelli G. “Nothing in nature that is not useful”. The anti-vaccination crusade and the idea of ‘harmonia naturae’ in Alfred Russel Wallace. Nuncius. 1992;7(1):109–130. https://doi.org/10.1163/182539192X00055.

14. Wolfe RM, Sharp LK. Anti-vaccinationists past and present. Bmj. 2002;325(7361):430–432. https://doi.org/10.1136/bmj.325.7361.430.

15. Aronowitz RA. The rise and fall of the lyme disease vaccines: a cautionary tale for risk interventions in American medicine and public health. The Milbank Quarterly. 2012;90(2):250–277. https://doi.org/10.1111/j.1468-0009.2012.00663.x.

16. Debus M, Tosun J. Political ideology and vaccination willingness: implications for policy design. Policy sciences. 2021;54(3):477–491. https://doi.org/10.1007/s11077-021-09428-0.

17. Joslyn MR, Sylvester SM. The determinants and consequences of accurate beliefs about childhood vaccinations. American Politics Research. 2019;47(3):628–649. https://doi.org/10.1177%2F1532673X17745342.

18. Daley MF, Narwaney KJ, Shoup JA, et al. Addressing parents’ vaccine concerns: a randomized trial of a social media intervention. American journal of preventive medicine. 2018;55(1):44–54. https://doi.org/10.1016/j.amepre.2018.04.010.

19. Fisher KA, Bloomstone SJ, Walder J, et al. Attitudes Toward a Potential SARS-CoV-2 Vaccine: a survey of US adults. Annals of internal medicine. 2020;173(12):964–973. https://doi.org/10.7326/M20-3569.

20. Lazarus JV, Ratzan SC, Palayew A, et al. A global survey of potential acceptance of a COVID-19 vaccine. Nature Medicine. 2021;27(2):225–228. https://doi.org/10.1038/s41591-020-1124-9.

21. Roozenbeek J, Schneider CR, Dryhurst S, et al. Susceptibility to misinformation about COVID-19 around the world. Royal Society open science. 2020;7(10):201199. https://doi.org/10.1098/rsos.201199.

22. Mohamed NA, Solehan HM, Mohd Rani MD, et al. Knowledge, acceptance and perception on COVID-19 vaccine among Malaysians: A web-based survey. Plos one. 2021;16(8):e0256110. https://doi.org/10.1371/journal.pone.0256110.

23. Bertin P, Nera K, Delouvée S. Conspiracy beliefs, rejection of vaccination, and support for hydroxychloroquine: A conceptual replication-extension in the COVID-19 pandemic context. Frontiers in psychology. 2020:2471. https://doi.org/10.3389/fpsyg.2020.565128.

24. Bradley VC, Kuriwaki S, Isakov M, et al. Unrepresentative big surveys significantly overestimated US vaccine uptake. Nature. 2021;600(7890):695–700. https://doi.org/10.1038/s41586-021-04198-4.

25. Nehal KR, Steendam LM, Ponce MC, et al. Worldwide vaccination willingness for COVID-19: A systematic review and meta-analysis. Vaccines. 2021;9(10). https://doi.org/10.3390/vaccines9101071.

26. Nindrea RD, Usman E, Katar Y, et al. Acceptance of COVID-19 vaccination and correlated variables among global populations: A systematic review and meta-analysis. Clinical Epidemiology and Global Health. 2021;12:100899. https://doi.org/10.1016/j.cegh.2021.100899.

27. Troiano G, Nardi A. Vaccine hesitancy in the era of COVID-19. Public Health. 2021;194:245–251. https://doi.org/10.1016/j.puhe.2021.02.025.

28. Barry M, Temsah MH, Alhuzaimi A, et al. COVID-19 vaccine confidence and hesitancy among health care workers: A cross-sectional survey from a MERS-CoV experienced nation. PLoS ONE. 2021;16(11):e0244415. https://doi.org/10.1371/journal.pone.0244415.

29. Biswas N, Mustapha, T, Khubchandani J, et al. The Nature and Extent of COVID-19 Vaccination Hesitancy in Healthcare Workers. Journal of Community Health. 2021:46(6):1244–1251. https://doi.org/10.1007/s10900-021-00984-3.

30. Lucia VC, Kelekar A, Afonso NM. COVID-19 vaccine hesitancy among medical students. Journal of Public Health. 2021;43(3):445–449. https://doi.org/10.1093/pubmed/fdaa230.

31. Li M, Luo Y, Watson R, et al. Healthcare workers’(HCWs) attitudes and related factors towards COVID-19 vaccination: A rapid systematic review. Postgraduate medical journal. 2021;0:1–7. http://dx.doi.org/10.1136/postgradmedj-2021-140195.

32. Skirrow H, Barnett S, Bell S, et al. Women’s views on accepting COVID-19 vaccination during and after pregnancy, and for their babies: a multi-methods study in the UK. BMC Pregnancy and Childbirth. 2022;22(1):1–15. https://doi.org/10.1186/s12884-021-04321-3.

33. Kaplan RM, Milstein A. Influence of a COVID-19 vaccine’s effectiveness and safety profile on vaccination acceptance. Proceedings of the National Academy of Sciences of the United States of America. 2021;118(10). https://doi.org/10.1073/pnas.2021726118.

34. Frankovic, K. Why won’t Americans get vaccinated? 2021. https://today.yougov.com/topics/politics/articles-reports/2021/07/15/why-wont-americans-get-vaccinated-poll-data. Accessed 18 Apr 2022.

35. Boseley, S. Fears of side-effects fuel reluctance to get COVID-19 jabs, survey finds. 2021. https://www.theguardian.com/society/2021/jun/04/fear-side-effects-fuel-covid-vaccine-reluctance-survey. Accessed 18 Apr 2022.

36. Shaw J, Stewart T, Anderson K B, et al. Assessment of US healthcare personnel attitudes towards coronavirus disease 2019 (COVID-19) vaccination in a large university healthcare system. Clinical Infectious Diseases. 2021;73(10):1776–1783.

37. Alabdulla M, Reagu SM, Al-Khal A, et al. COVID-19 vaccine hesitancy and attitudes in Qatar: A national cross-sectional survey of a migrant-majority population. Influenza Other Respi Viruses. 2021;15:361–370. https://doi.org/10.1111/irv.12847.

38. Gerretsen P, Kim J, Caravaggio F, et al. Individual determinants of COVID-19 vaccine hesitancy. PloS one. 2021;16(11):e0258462. https://doi.org/10.1371/journal.pone.0258462.

39. Cerda, AA, García LY. Hesitation and refusal factors in individuals’ decision-making processes regarding a coronavirus disease 2019 vaccination. Frontiers in public health. 2021;9. https://doi.org/10.3389/fpubh.2021.626852.

40. Shih SF, Wagner AL, Masters NB, et al. Vaccine Hesitancy and Rejection of a Vaccine for the Novel Coronavirus in the United States. Frontiers in Immunology. 2021;12:1–8. https://doi.org/10.3389/fimmu.2021.558270.

41. Reiter PL, Pennell ML, Katz ML. Acceptability of a COVID-19 vaccine among adults in the United States: How many people would get vaccinated?. Vaccine. 2020;38(42):6500–6507. https://doi.org/10.1016/j.vaccine.2020.08.

42. Zhang X, Guo Y, Zhou Q, et al. The mediating roles of medical mistrust, knowledge, confidence and complacency of vaccines in the pathways from conspiracy beliefs to vaccine hesitancy. Vaccines. 2021;9(11):1–15. https://doi.org/10.3390/vaccines9111342.

43. Lin C, Tu P, Beitsch L. Confidence and Receptivity for COVID-19 Vaccines: A Rapid Systematic Review. Vaccines. 2021;9(1):16. https://doi.org/10.3390/vaccines9010016.

44. Ruiz JB, Bell R. Predictors of intention to vaccinate against COVID-19: Results of a nationwide survey. Vaccine. 2021;39(7):1080–1086. https://doi.org/10.1016/j.vaccine.2021.01.010.

45. Schwarzinger M, Watson V, Arwidson P, et al. COVID-19 vaccine hesitancy in a representative working-age population in France: a survey experiment based on vaccine characteristics. The Lancet Public Health. 2021;6(4):e210–e221. https://doi.org/10.1016/S2468-2667(21)00012-8.

46. Bendau A, Plag J, Petzold M, et al. COVID-19 vaccine hesitancy and related fears and anxiety. International Immunopharmacology. 2021;97:107724. https://doi.org/10.1016/j.intimp.2021.107724.

47. Akesson J, Ashworth-Hayes S, Hahn R, et al. Fatalism, Beliefs, and Behaviors during the COVID-19 Pandemic. 2020. Working Paper No. 27245. NBER. https://doi.org/10.3386/w27245.

48. Hou Z, Tong Y, Du F, et al. Assessing COVID-19 vaccine hesitancy, confidence, and public engagement: A global social listening study. Journal of medical Internet research. 2021;23(6):e27632. https://doi.org/10.2196/27632.

49. De Figueiredo A, Larson HJ, Reicher SD. The potential impact of vaccine passports on inclination to accept COVID-19 vaccinations in the United Kingdom: Evidence from a large cross-sectional survey and modeling study. EClinicalMedicine. 2021;40:101109. https://doi.org/10.1016/j.eclinm.2021.101109.

50. Mills MC, Rüttenauer T. The effect of mandatory COVID-19 certificates on vaccine uptake: synthetic-control modelling of six countries. The Lancet Public Health. 2022;7(1):e15–e22. https://doi.org/10.1016/S2468-2667(21)00273-5.

51. Oliu M, Pradelski BSR, Woloszko N, et al. The Effect of COVID-19 Certificates on Vaccine Uptake, Health Outcomes, and the Economy. Bruegel Working Paper, 01/2022. 2022;1–24. https://doi.org/10.21203/rs.3.rs-1242919/v1.

52. Jennings W, Stoker G, Bunting H, et al. Lack of trust, conspiracy beliefs, and social media use predict COVID-19 vaccine hesitancy. Vaccines. 2021;9(6):593. https://doi.org/10.3390/vaccines9060593.

53. Centers for Disease Control and Prevention. Myths and Facts about COVID-19 Vaccines. 2021. https://www.cdc.gov/coronavirus/2019-ncov/vaccines/facts.html. Accessed 18 Apr 2022.

54. European Commission. Questions and answers on COVID-19 vaccination in the EU. https://ec.europa.eu/info/live-work-travel-eu/coronavirus-response/safe-COVID-19-vaccines-europeans/questions-and-answers-COVID-19-vaccination-eu_en. Accessed 18 Apr 2022.

55. Baumgaertner B, Carlisle JE, Justwan F. The influence of political ideology and trust on willingness to vaccinate. PLoS ONE. 2018;13(1):1–13. https://doi.org/10.1371/journal.pone.0191728.

56. Tyson, A. Republicans remain far less likely than Democrats to view COVID-19 as a major threat to public health. 2020. https://www.pewresearch.org/fact-tank/2020/07/22/republicans-remain-far-less-likely-than-democrats-to-view-covid-19-as-a-major-threat-to-public-health/. Accessed 18 Apr 2022.

57. Latkin CA, Dayton L, Yi G, et al. Trust in a COVID-19 vaccine in the U.S.: A social-ecological perspective. Social Science and Medicine. 2021;270:113684. https://doi.org/10.1016/j.socscimed.2021.113684.

58. Summers J. Timeline: How Trump Has Downplayed The Coronavirus Pandemic. 2020. https://www.npr.org/sections/latest-updates-trump-COVID-19-results/2020/10/02/919432383/how-trump-has-downplayed-the-coronavirus-pandemic. Accessed 18 Apr 2022.

59. Saad L. More in U.S. Vaccinated After Delta Surge, FDA Decision. 2021. https://news.gallup.com/poll/355073/vaccinated-delta-surge-fda-decision.aspx. Accessed 18 Apr 2022.

60. Cornelson K, Miloucheva B. Political polarization, social fragmentation, and cooperation during a pandemic. 2020. University of Toronto, Department of Economics.

61. Cowan SK, Mark N, Reich JA COVID-19 vaccine hesitancy is the new terrain for political division among Americans. Socius: Sociological Research for a Dynamic World. 2021;7:1–3.

62. Fine P, Eames K, Heymann DL. “Herd immunity”: a rough guide. Clinical Infectious Diseases, 2011;52(7):911–916. https://doi.org/10.1093/cid/cir007.

63. Aschwanden C. Five reasons why COVID herd immunity is probably impossible. Nature. 2021;591:520–522.

64. Fontanet A, Cauchemez S. COVID-19 herd immunity: where are we?. Nature Reviews Immunology. 2020;20(10):583–584. https://doi.org/10.1038/s41577-020-00451-5.

65. Frederiksen LSF, Zhang Y, Foged, C, et al. The long road toward COVID-19 herd immunity: vaccine platform technologies and mass immunization strategies. Frontiers in immunology, 2020;11:1817. https://doi.org/10.3389/fimmu.2020.01817.

66. Randolph HE, Barreiro LB. Herd immunity: understanding COVID-19. Immunity. 2020;52(5): 737–741. https://doi.org/10.1016/j.immuni.2020.04.012.

67. Jonsen AR. “The ethics of organ transplantation: A brief history”. Virtual Mentor. 2012;14 (3):264–268.

68. Ritchie H, et al. Coronavirus pandemic (COVID-19). Our world in data. 2020. https://ourworldindata.org/covid-vaccinations?country=OWID_WRL. Accessed 18 May 2022.

69. Van der Veen O. Political Polarisation Compared: Creating a Comprehensive Index of Political Polarisation. Doctoral dissertation, Central European University. 2021.

70. Mathieu E, et al. A global database of COVID-19 vaccinations. Nature human behaviour. 2021; 5(i7);947–953. https://doi.org/10.1038/s41562-021-01122-8.

71. Berniell I, Fawaz Y, Laferrère A, et al. The COVID-19 curtain: can past communist regimes explain the vaccination divide in Europe?. http://sedici.unlp.edu.ar/bitstream/handle/10915/128344/Documento_completo.pdf-PDFA.pdf?sequence=1. Accessed 18 May 2022.

72. Kennedy, J. Populist politics and vaccine hesitancy in Western Europe: an analysis of national-level data. European journal of public health. 2019;29(3):512–516. https://doi.org/10.1093/eurpub/ckz004.

73. Eurostat (2022), Database. Deaths by week, sex and NUTS 2 region https://ec.europa.eu/eurostat/databrowser/view/DEMO_R_MWK2_TScustom_2533461/default/table?lang=en. Accessed 19 Apr 2022.

